# Rapid Molecular Detection of SARS-CoV-2 (COVID-19) Virus RNA Using Colorimetric LAMP

**DOI:** 10.1101/2020.02.26.20028373

**Authors:** Yinhua Zhang, Nelson Odiwuor, Jin Xiong, Luo Sun, Raphael Ohuru Nyaruaba, Hongping Wei, Nathan A. Tanner

## Abstract

The ability to detect an infectious agent in a widespread epidemic is crucial to the success of quarantine efforts in addition to sensitive and accurate screening of potential cases of infection from patients in a clinical setting. Enabling testing outside of sophisticated laboratories broadens the scope of control and surveillance efforts, but also requires robust and simple methods that can be used without expensive instrumentation. Here we report a method to identify SARS-CoV-2 (COVID-19) virus RNA from purified RNA or cell lysis using loop-mediated isothermal amplification (LAMP) using a visual, colorimetric detection. This test was additionally verified using RNA samples purified from respiratory swabs collected from COVID-19 patients in Wuhan, China with equivalent performance to a commercial RT-qPCR test while requiring only heating and visual inspection. This simple and sensitive method provides an opportunity to facilitate virus detection in the field without a requirement for complex diagnostic infrastructure.

## INTRODUCTION

The emergence of a new coronavirus (2019-nCoV, now named SARS-CoV-2) has infected tens of thousands of people in China, with cases in at least 28 other countries and prompting a worldwide response. The diagnostics industry has responded rapidly, with Emergency Use Authorization granted for PCR-based tests from the US CDC (https://www.cdc.gov/coronavirus/2019-ncov/about/testing.html), Seegene in Korea and BGI in China (https://www.bgi.com/global/company/news/bgi-develops-real-time-dna-based-kit-for-detecting-the-2019-novel-coronavirus/) among many other providers releasing reagents, primers and probes, and other materials to support this urgent public health need. The current diagnostic standard combines clinical symptoms and molecular method, and as many of the symptoms resembles those of common cold and influenza, an accurate molecular result is critical for final diagnosis. These molecular methods include metagenomics sequencing mNGS (1) and RT-qPCR(2), both are excellent and sensitive techniques, but approaches not without limitations. mNGS is restricted by throughput, turnover time, formidable high cost and requirement for high technical expertise. As with most molecular diagnostics, RT-qPCR is the most widely used method, but it requires expensive laboratory instruments and is difficult to utilize outside of well-equipped facilities. In combination to the different patient samples containing variable number of virus, a high proportion of patients were diagnosed as false negatives (3,4).

Here we describe a molecular diagnostic approach for SARS-CoV-2 RNA detection using loop-mediated isothermal amplification (LAMP) and simple visual detection of amplification for potential use in rapid, field applications. LAMP was developed as a rapid and reliable method to amplify from a small amount target sequence at a single reaction temperature, obviating the need for sophisticated thermal cycling equipment (5). Since the initial description of LAMP, a number of advancements in detection technology have helped establish LAMP as a standard method for simple isothermal diagnostics. These detection methods have allowed detection by visual examination without instrumentation using dyes that utilize inherent by-products of the extensive DNA synthesis, such as malachite green(6), calcein (7) and hydroxynaphthol blue (8). We recently developed a method for visual detection of LAMP amplification (9) using pH-sensitive dyes to exploit the change of pH that resulting from proton accumulation due to dNTP incorporation. This method has been used in a large scale field survey of Wolbachia-containing mosquitos (10), Grapevine red blotch virus without DNA extraction (11), testing urine samples for Zika virus (12) and even amplification detection on the International Space Station (13). The breadth of application highlights the applicability of visual detection methods to provide an advantage in simplicity and portability for enabling new, rapid diagnostics.

This study describes testing and validation of 5 sets LAMP primers targeting two fragments of the SARS-CoV-2 genome using short (∼300bp) RNA fragments made with *in vitro* transcription and RNA samples from patients. We also demonstrate compatibility with simple approaches to RNA purification to simplify the detection procedure and avoid complex RNA extraction, and with clinical swab samples taken from COVID-19 patients. Our aim is to share this information in order to help develop a reliable and easy method to detect this viral RNA outside of sophisticated diagnostic laboratories and expand the toolbox of molecular tests used to combat and surveil this growing public health threat.

## MATERIALS AND METHODS

### LAMP Primer Design and Testing

We designed 5 sets of LAMP primers targeting two fragments (Table 1) of SARS-CoV-2 sequence (GenBank accession number MN908947) using the online software Primer Explorer V5 (available for free use at: https://primerexplorer.jp/e/). DNA fragments containing these two regions were synthesized as gBlocks (Integrated DNA Technologies) and T7 RNA polymerase promoters were added by PCR (NEB M0493, numbers indicate NEB catalog ID unless otherwise noted) using primer pairs where one primer containing the promoter sequence. RNAs were then synthesized by *in vitro* transcription (E2050) using these PCR products as templates and purified using RNA clean up columns (T2040). The resulting RNAs as well as the gBlocks were serially diluted in 10-fold increments using 0.1x TE buffer containing 0.01% Tween 20. RT-LAMP reactions were performed using WarmStart® Colorimetric LAMP 2X Master Mix (DNA & RNA) (M1800) supplemented with 1 μM SYTO®-9 double-stranded DNA binding dye (Thermo Fisher S34854) and incubated on a real-time qPCR machine (BioRad CFX96) for 120 cycles with 15 seconds each cycle (total ∼40 min). The color of the finished reactions was recorded using an office flatbed scanner. Synthetic RNAs were spiked into Hela cells, which were then diluted and lysed using Luna® Cell Ready Lysis Module (E3032). Each lysate was then diluted 10x with 0.1x TE +0.01% Tween 20 and 1 μL was added to standard colorimetric LAMP reactions. For compatibility with blood recovery, RNA was spiked into 200 μL whole human blood (Quadrant Health Strategies) and then purified the total blood RNA using Monarch® Total RNA Miniprep Kit (T2010).

**Table 1.**
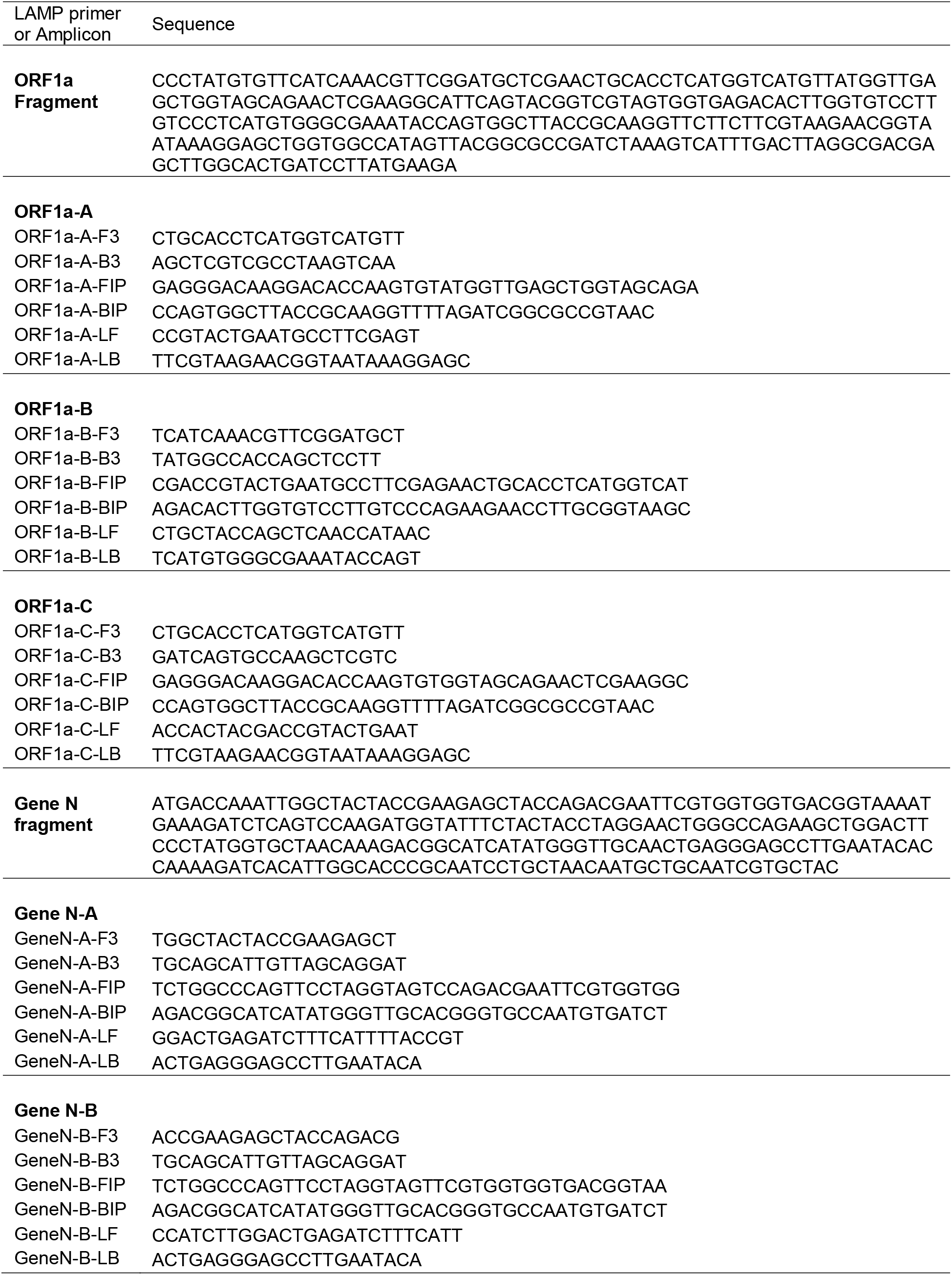
Sequences of amplicons and LAMP primers

### Sample Collection and Processing

Wuhan Institute of Virology CAS is one of the authorized labs approved by CDC of Wuhan city for detecting COVID-19 virus in clinical samples. All samples were handled and deactivated first in biosafety level 2 laboratory with personal protection equipment for biosafety level 3 following the guidelines for detecting nucleic acid of SARS-CoV-2 in clinical samples. Research on developing new diagnostic techniques for COVID-19 using clinical samples has been approved by the ethical committee of Wuhan Institute of Virology. Briefly, respiratory specimens (swabs) collected from patients admitted to various Wuhan health care facilities were immediately placed into sterile tubes containing 3ml of viral transport media (VTM). The medium constitutes Hank’s balanced salt solution at pH 7.4 containing BSA (1%), amphotericin (15 μg/mL), penicillin G (100 units/mL), and streptomycin (50 μg/mL). The swabs were deactivated by heating at 56 °C for 30 minutes in a biosafety level 2 (BSL 2) laboratory at the Wuhan Institute of Virology in Zhengdian Park. Due to the possibility of generating aerosols, the samples were allowed to cool at room temperature before further processing. If the samples were not for use immediately, they were stored at 4 °C.

### Extraction of Viral RNA and RT-PCR detection

Total viral RNA was extracted from the deactivated samples using automated extraction instrument, Purifier™ Modesty (Genfine Biotech, Beijing-China, LTD), according to the manufacturer’s instructions. The instrument is capable of processing 32 samples per run for 21 minutes with less hands-on work. Furthermore, it is equipped with a UV light lamp to ensure biosafety of the user. A commercial COVID-19 RT-PCR kit (Jienuo Inc, Shanghai, China) was used to detect if the samples are positive or negative to COVID-19.

### LAMP Assays on RNA Samples From COVID-19 Patients

The assay was performed in a 20 µl reaction mixture containing 2 µL of 10x primer mix of 16 µM (each) of Forward Inner Primer (FIP) and Backward Inner Primer (BIP), 2 µM (each) of F3 and B3 primers, 4 µM (each) of Forward Loop (LF) and Backward Loop (LB) primers, 10 µL of WarmStart Colorimetric Lamp 2X Master Mix (M1800) 5 µL of DNAse, RNAase free water (Beyotime Biotech, China),and 3 µl of RNA template. The reaction mixture was set at 65 °C for 30 minutes on a dry bath (Avist Technology Co., Wuhan, China).

## RESULTS AND DISCUSSION

We designed 5 full LAMP primers sets targeting SARS-CoV-2 RNA, with amplicon regions designed to the 5′ region of the ORF1a gene and Gene N. Each set was tested with synthetic DNA substrates and RNA transcribed from that DNA substrate before consideration for use clinically. To evaluate detection sensitivity, synthetic RNAs were serially diluted from ∼120 million copies down to ∼120 copies (per 25 μL reaction) at 10-fold intervals in LAMP reactions. All five primer sets showed similar detection sensitivity and could consistently detect as low as a few hundred copies, with sporadic detection of 120 copies (or 4.8 copies/μL; Fig. 1). The results from colorimetric detection were 100% in agreement with the real time detection. To estimate the relative efficiency using RNA or DNA templates, we compared synthetic RNA with similarly diluted gBlock dsDNA on real-time LAMP signal (Fig. 2). For the 2 primer sets we compared, one showed slightly slower amplification and detection with RNA template while the other appeared slightly faster, confirming the RNA is efficiently converted to cDNA by the reverse transcriptase (WarmStart RTx) and subsequently amplified via LAMP by the DNA-dependent DNA polymerase (*Bst* 2.0 WarmStart).

**Figure 1.**
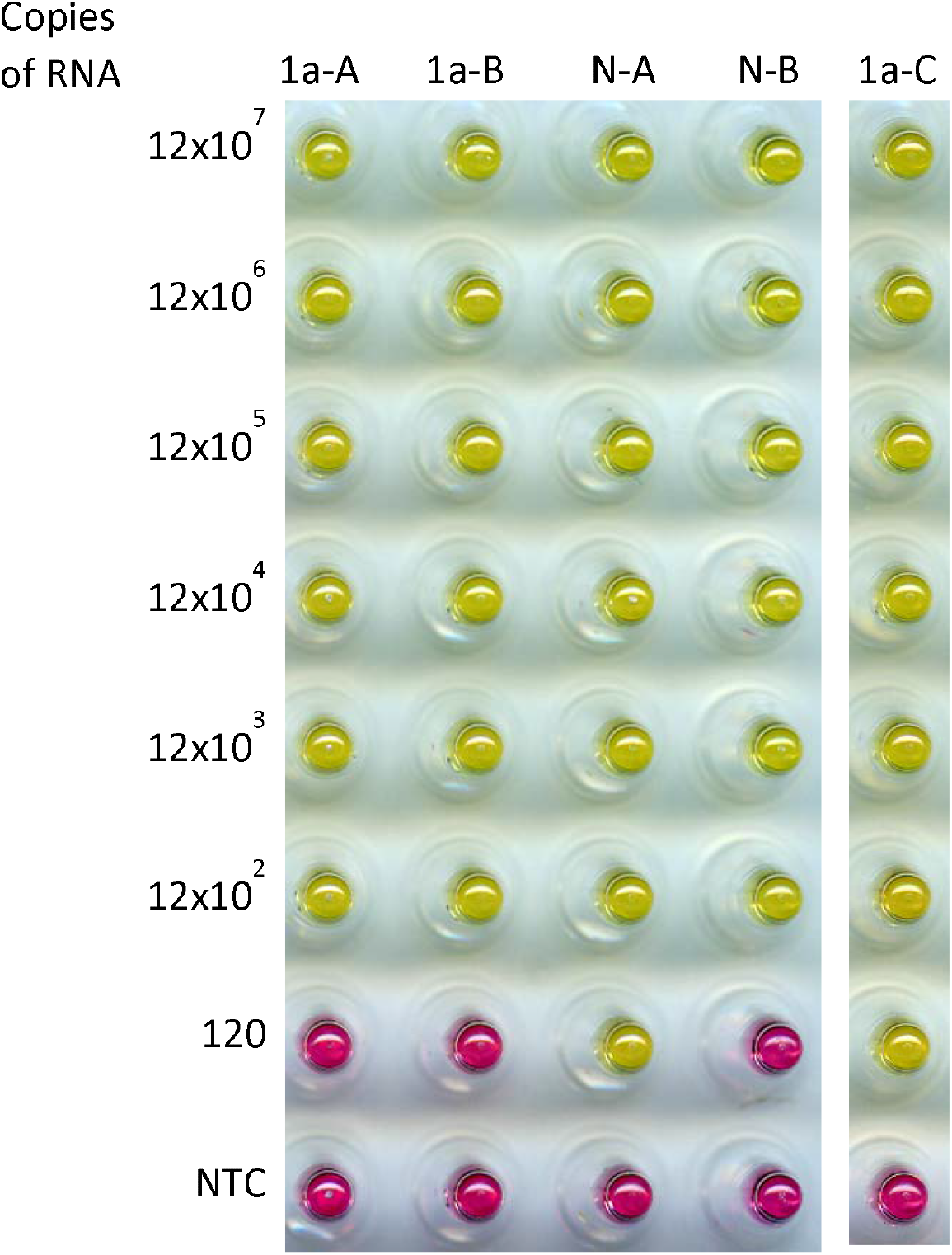
Detection sensitivity of synthetic 2019-nCoV RNA amplicons. 5 sets of LAMP primers were tested with templates ranging from 120×10^6^ to 120 copies. Yellow, positive amplification; Pink, no amplification.

**Figure 2.**
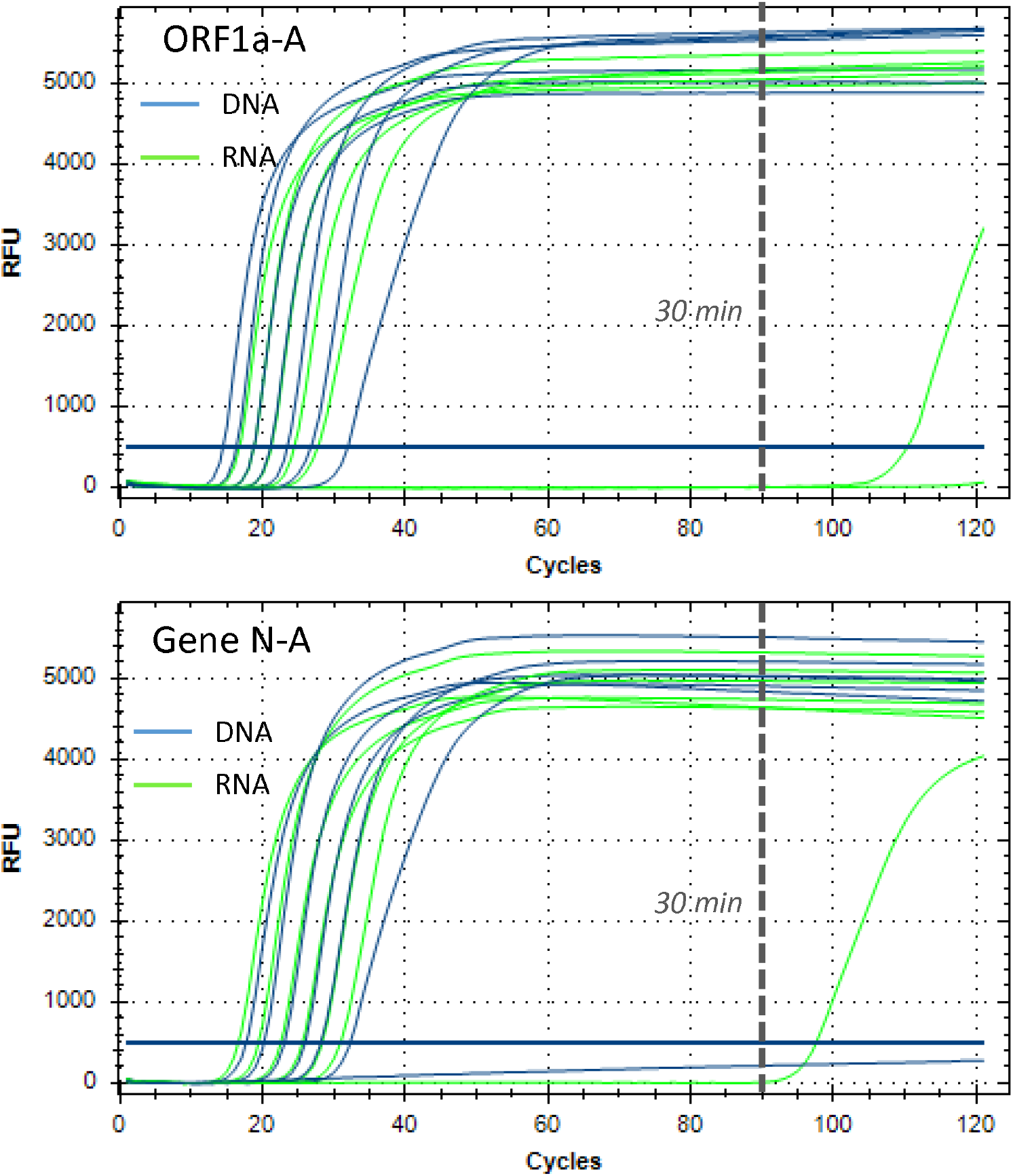
Comparison of RNA and gBlock dsDNA templates in LAMP amplification using real time amplification curves. Two primer sets (ORF1a-A and Gene N-A) were used to amplify either RNA (green curves, dilutions from 120 ×10^6^ to 120 copies) or gDNA (blue curves, 60×10^6^ to 60 copies). For ORF1a-A primer set, the gBlock is faster than RNA template; For Gene N-A primer set, the RNA is slightly faster. Each “cycle” represents 20 seconds, with 30 minute timepoint noted by dashed line.

With current diagnostic methods, e.g. RT-qPCR, purified RNA is used in the input. This will of course enable maximum efficiency and sensitivity but requires skill and instrumentation in addition to adding extra time to the diagnostic workflow. We investigated whether it is possible to perform detection using crude cell lysate in order to avoid this RNA purification step. The results indicated that ∼480 copies were detected with all four primer sets, a similar sensitivity as the detection sensitivity with synthetic RNA alone (Fig. 3A) with no interference by the lysate to either the amplification efficiency or visual color change. We also tested whether we could recover the synthetic RNA spiked into biological sample with a mock experiment during purification of total RNA. We spiked various amount of synthetic RNA into whole human blood and purified the total blood RNA. We were able to recover and detect the spiked RNA (Fig. 3B), indicating the total RNA has no interference during the purification or the detection process. While the column-based approach is less compatible with the simple, field detection enabled by colorimetric LAMP, this is a typical laboratory workflow and can be used with simple isothermal amplification in a similar fashion to more expensive and involved qPCR detection workflows.

**Figure 3.**
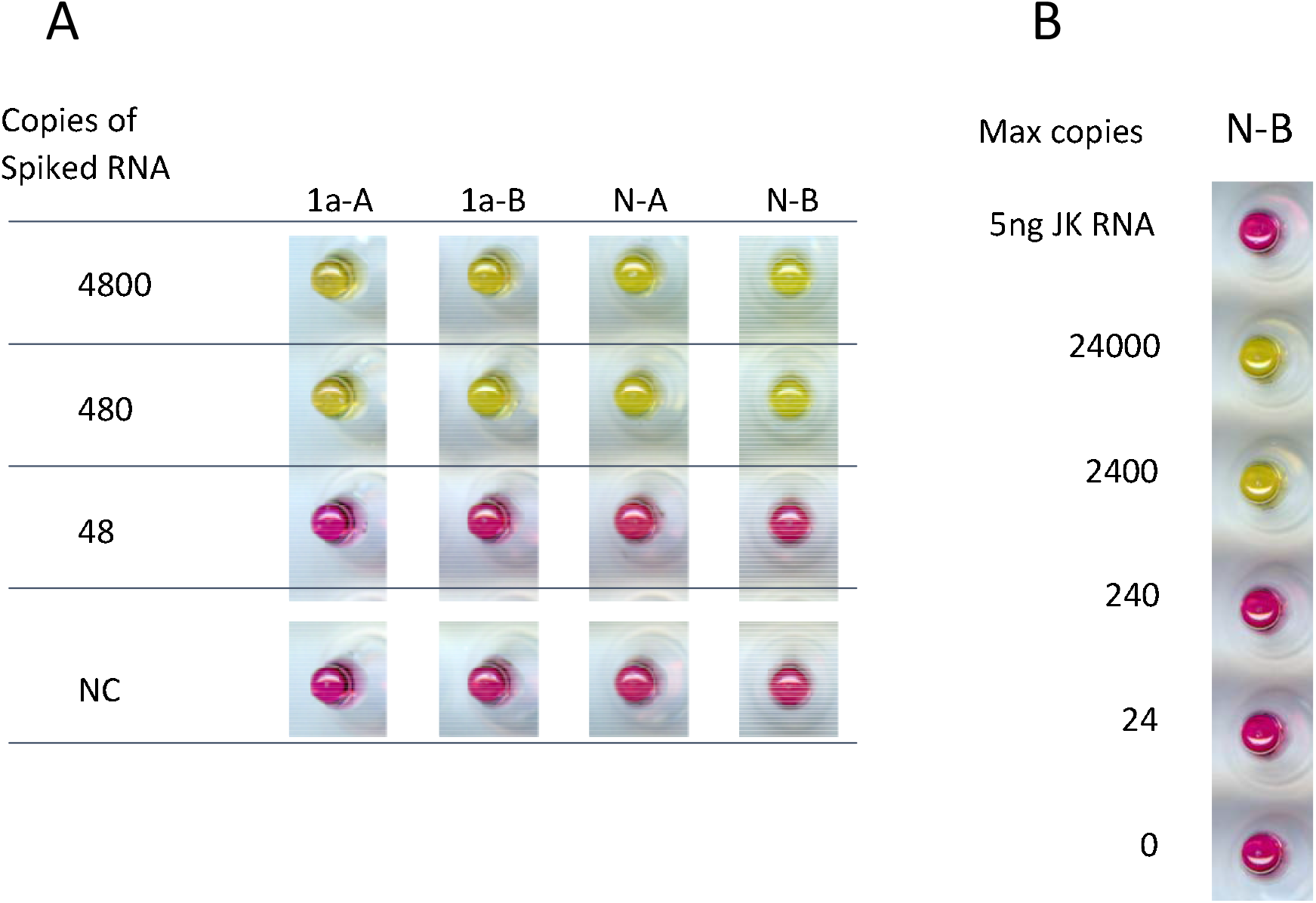
**A**. Direct RNA LAMP detection using total cell lysate. The approximate maximum number of copies of synthetic RNA added to each LAMP reaction are shown. For 4800 copies, there were about 200 Hela cells present. NC, no cell and no template control. **B**. LAMP detection of target RNA spiked in whole blood. The maximal possible copies of the target RNA that could be recovered were shown. In a control, 5ng of Jurkat total RNA was added, which is similar to the total RNA present in the reaction with blood samples.

With this preliminary evaluation of potential LAMP primer sets, the best performing sets (ORF1a-A, GeneN-A) were shared and synthesized in Wuhan for testing with actual COVID-19 samples. RNA was extracted from swabs using the standard laboratory protocol (see Materials and Methods) and RNA tested in colorimetric RT-LAMP alongside a commercial RT-qPCR assay. In addition to controls, a total of 7 patient RNA samples were tested, 6 of which were determined to be positive by RT-qPCR using ORF1a primers (C_q_ 25–36.5, Table 2) and 4 positive with Gene N primers. One sample was negative by both RT-qPCR primer sets. When these samples were tested in the colorimetric LAMP assay, all 6 RT-qPCR positive samples showed visible color change indicating positive amplification, while the single RT-qPCR negative samples maintained pink color and was judged negative (Figure 4). Thus the colorimetric LAMP assay showed 100% agreement with the RT-qPCR results across a range of C_q_ values. Although a small number of samples were tested here, the colorimetric LAMP assay enables reliable SARS-CoV-2 detection without sophisticated instrumentation, matching the RT-qPCR performance in field and point-of-care settings.

**Table 2.** Comparison of detection accuracy between RT-LAMP and RT-PCR with COVID-19 patient samples

| Sample ID | RT-LAMP |  | RT-qPCR (C<sub>q</sub>) |  |
| --- | --- | --- | --- | --- |
|  | ORF1a | N | ORF1a | N |
| 1 | + | + | 25.74 | 21.15 |
| 2 | + | + | 33.61 | 28.65 |
| 3 | + | + | 34.26 | 28.68 |
| 4 | + | + | 36.26 | N/A |
| 5 | + | + | 25.30 | 30.25 |
| 6 | + | + | 36.51 | N/A |
| 7 | - | - | N/A | N/A |
| Blank<sup>a</sup> | - | - | N/A | N/A |
| Negative Control | - | - | N/A | N/A |
| Positive Control<sup>b</sup> | - | - | 24.46 | N/A |

+ Positive reaction; - Negative reaction; N/A not detected
<sup>a</sup> No template control; <sup>b</sup> plasmid DNA (standard control) for the RT-PCR kit

**Figure 4.**
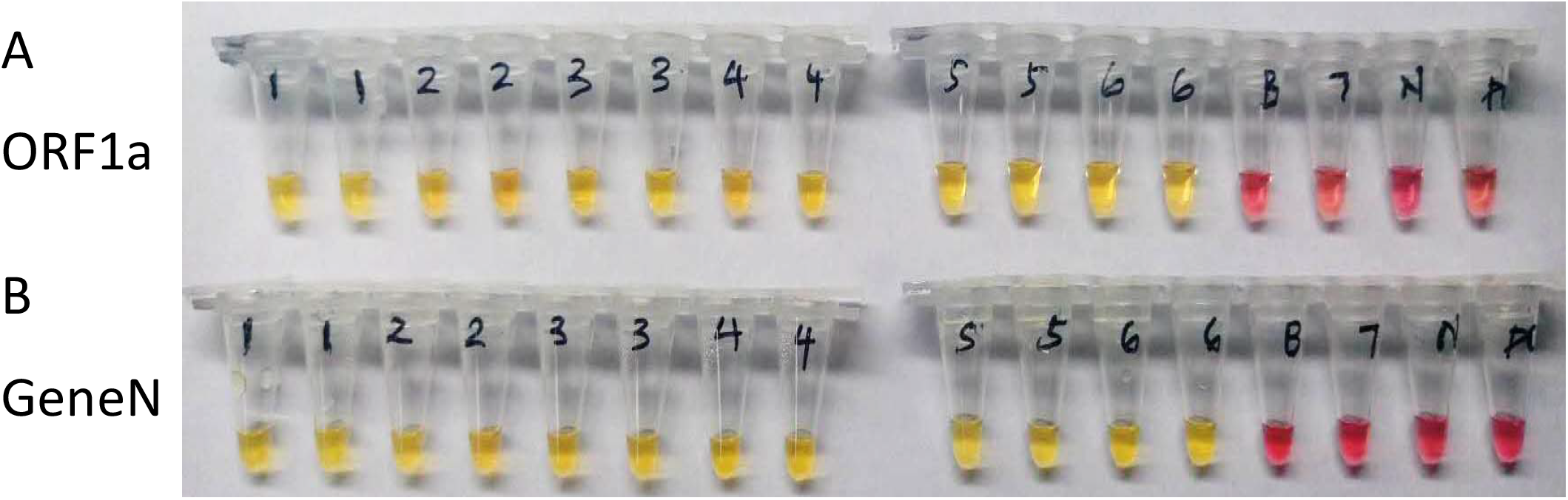
SARS-CoV-2 detection from COVID-19 patient samples in Wuhan, China. Samples testing positive (1-6) or negative (7) with commercial RT-qPCR tests were assayed using colorimetric LAMP assay with primer set targeting ORF1a (A) and GeneN (B). Yellow indicates a positive detection after 30 min incubation, and pink a negative reaction with results compared to the negative control (N). B, Blank control without template. P, samples containing the plasmid used as positive control for qPCR.

In conclusion, colorimetric LAMP provides a simple, rapid method for SARS-CoV-2 RNA detection. Not only purified RNA can be used as the sample input, but also direct tissue or cell lysate may be used without an RNA purification step. This combination of a quick sample preparation method with an easy detection process may allow the development of portable, field detection in addition to a rapid screening for point-of-need testing applications. This virus represents an emerging significant public health concern and expanding the scope of diagnostic utility to applications outside of traditional laboratories will enable greater prevention and surveillance approaches. The efforts made here will serve as a model for inevitable future outbreaks where the use of next generation portable diagnostics will dramatically expand the reach of our testing capabilities for better healthcare outcomes.

## Data Availability

No data is required other than what is contained in the manuscript

## ACKNOWLEDGEMENTS

We thank Dr. Guoping Ren for advice and assistance using the Luna Cell Ready Lysis Module.

